# Detecting a History of Repetitive Head Impacts from a Short Voice Recording

**DOI:** 10.1101/2021.09.20.21263753

**Authors:** Michael G. Tauro, Mirco Ravanelli, Cristian A. Droppelmann

**Affiliations:** ReLup, Toronto, Ontario, Canada; Mila, University of Montreal. Montreal, Quebec, Canada; Molecular Brain Research Group, Robarts Research Institute, University of Western Ontario, London, ON, Canada

**Keywords:** Chronic traumatic encephalopathy, Repetitive head impacts, Machine learning, Speech, Deep learning, Neurodegeneration, Transfer learning

## Abstract

Repetitive head impacts (RHI) are associated with an increased risk of developing various neurodegenerative disorders, such as Alzheimer’s disease (AD), Parkinson’s disease (PD), and most notably, chronic traumatic encephalopathy (CTE). While the clinical presentation of AD and PD is well established, CTE can only be diagnosed post-mortem. Therefore, a distinction can be made between the pathologically defined CTE and RHI-related functional or structural brain changes (RHI-BC) which may result in CTE. Unfortunately, there are currently no accepted biomarkers of CTE nor RHI-BC, a major hurdle to achieving clinical diagnoses. Interestingly, speech has shown promise as a potential biomarker of both AD and PD, being used to accurately classify individuals with AD and PD from those without. Given the overlapping symptoms between CTE, RHI-BC, PD and AD, we aimed to determine if speech could be used to identify individuals with a history of RHI from those without. We therefore created the Verus dataset, consisting of 13 second voice recordings from 605 professional fighters (RHI group) and 605 professional athletes in non-contact sports (control group) for a total of 1210 recordings. Using a deep learning approach, we achieved 85% accuracy in detecting individuals with a history of RHI from those without. We then used our model trained on the Verus dataset to fine-tune on publicly available AD and PD speech datasets and achieved new state-of-the-art accuracies of 84.99% on the AD dataset and 89% on the PD dataset. Finding a biomarker of CTE and RHI-BC that presents early in disease progression is critical to improve risk management and patient outcome. Our study is the first we are aware of to investigate speech as such a candidate biomarker of RHI-BC.

## 1. Introduction

Concussion is defined as a form of mild traumatic brain injury (mTBI) that disrupts regular brain function [71]. Concussion can be caused by blunt force trauma to the head or rapid acceleration of the head, as occurs in whiplash [77]. While the acute symptoms of concussion generally resolve within a week, long term effects can occur from multiple or even a single concussion [52]. However, focusing on the long-term risks associated exclusively with concussion may miss key information. Athletes for example tend to underreport the number of concussions they have had, making analyses between health outcomes and number of concussions difficult. Further, there is no objective threshold one can use to differentiate a concussion from a subconcussive blow to the head [52]. Finally, recent literature suggests that the behavioural symptoms of concussion result from physical disruption to brain function, while subconcussive impacts have the potential to cause the long-term effects often attributed to concussion [81]. Therefore, we will use the term repetitive head impacts (RHI) in reference to blows to the head which may or may not have resulted in a clinically diagnosed concussion. Correspondingly, a history of RHI is associated with an increased risk of developing Alzheimer’s disease (AD), Parkinson’s disease (PD), amyotrophic lateral sclerosis (ALS), chronic traumatic encephalopathy (CTE) and other disabilities [43, 34, 76, 13, 42].

CTE is a neurodegenerative disorder linked to a history of RHI [55]. Originally documented in professional boxers, CTE has now been identified in former professional football, hockey, soccer and rugby players, as well as combat veterans and others [69, 16, 61, 50]. While the symptoms of CTE are quite heterogeneous, [55] has proposed a staged-based characterisation of the disease, with increasing stages relating to increased neurodegeneration and symptom severity. Stage 1 may be asymptomatic, with memory problems occurring rarely. Stage 2 symptoms include severe depressive episodes as well as behavioural changes and outbursts. Stage 3 is characterized by the first signs of cognitive deficits, such as executive dysfunction, memory loss, attentional and concentration difficulties and explosive outbursts. Stage 4 symptoms may include more severe cognitive deficits, including increased deficits in memory, attention, executive functioning, language, as well as severe depression, suicidal tendencies, paranoia, gait and visuospatial difficulties, dysarthria and parkinsonism.

CTE is pathologically characterized, with diagnoses only possible post-mortem [54]. Indeed, the clinical presentation of CTE is not universally agreed upon, nor is the number or severity of RHI needed to precipitate CTE [82, 43, 30]. Therefore, in this paper a distinction is made between CTE, and RHI-related functional or structural brain changes (RHI-BC). Unfortunately, there are no accepted biomarkers of CTE or RHI-BC [9]. As CTE is a degenerative disorder, early detection through the use of biomarkers could be critical in improving patient outcome and slowing disease progression.

The use of speech as a biomarker of neurodegenerative disorders has recently come under investigation with promising results [68]. Using voice recordings, [59] achieved 90% accuracy in classifying those with Parkinson’s disease (PD) from those without. [85] achieved 91.6% accuracy in classifying those with in classifying ALS patients with bulbar-onset from those without. While the previous two studies relied solely on the acoustic features of speech, [29] used a combination of acoustic, semantic, lexical, as well as other linguistic features to achieve 81.9% accuracy in classifying those with Alzheimer’s disease (AD) from those without. This warrants the investigation of speech as a biomarker of RHI-BC. In fact, not only does exposure to RHI increase the risk of developing PD, AD and ALS, but CTE also has overlapping symptoms with each of the aforementioned neurodegenerative disorders [55, 32]. Further, speech problems have long been considered a symptom of CTE. Indeed, [51] commented that many boxers examined had “hesitancy of speech”, “indistinct”, slow, and “thick, muffled, and hard to understand” speech, along with other issues. [23] described case reports of 11 boxers, with 6 listed as having motor speech problems. More recently, [46] characterised the speech of 102 active and retired professional fighters in comparison to 27 healthy controls. Professional fighters were reported to have a significantly slower articulation rate, with 88% of fighters having a slower articulation rate than the mean of the control group. Fighters also exhibited increased interruptions in speech, such as pauses, stuttering, and other dysfluencies. The authors noted that the speech characteristics of the fighters are also observed in several types of dysarthria, PD and parkinsonism. Yet, as speech problems are associated with RHI, and speech has been used to successfully detect neurodegenerative disorders that present similarly to CTE, speech might also serve as a biomarker of RHI-BC.

The investigation of speech as a biomarker of various diseases has historically been influenced by other popular speech classification tasks such as speaker recognition [68]. Traditionally, the best results in speaker recognition were achieved using some form or combination of a Gaussian Mixture Model (GMM) [31]. More specifically, a GMM-Universal Background Model (UBM) was used by [3] to predict PD severity in a longitudinal study. Yet the current trend has now shifted to the use of deep neural networks (DNN) [6]. Indeed, many recent performance advancements in speaker recognition and verification tasks are achieved through the use of x-vectors and other similar embedding approaches [73, 10]. x-vectors are fixed-length embedding vectors of variable length voice recordings. x-vectors have been used to achieve state of the art accuracy in detecting gender, language and PD. They have also recently been used to detect AD [59, 72, 66, 63]. Notably, x-vectors result in text-independent speaker identification, meaning x-vectors capture the acoustic properties of speech, but not the individual words or meaning. This was considered advantageous as motor-speech disorders occur earlier than language impairments in PD and ALS, thus we reasoned this could be the case in individuals exposed to RHI [36, 41, 83]. Thus, as our speaker recognition model, we decided to use the x-vector scheme with an Emphasized Channel Attention, Propagation and Aggregation-Time Delay Neural Network (ECAPA-TDNN) architecture. The ECAPA-TDNN model was developed by [27] and improved upon the previous TDNN-x-vector architecture to achieve state of the art performance in speaker recognition tasks.

The prevalence of CTE within populations exposed to RHI is unknown, with estimates ranging from 5-99% [18, 11, 57]. As the incidence rate and clinical presentation of CTE remain ambiguous, we aimed to determine if state of the art speech classification algorithms based on modern deep learning techniques could differentiate individuals with RHI exposure from those without. To do so, we created the Verus corpus, composed of 1210 individuals, corresponding to one 13 second voice recording each. Of these 1210 individuals, 605 were retired and active professional fighters and 605 were retired and active professional athletes in non-contact sports. The fighters were composed of boxers, mixed martial artists (MMA), kickboxers and Muay Thai fighters. These fighters were considered the group with RHI exposure, or the RHI group. The athletes in non-contact sports, considered the control group, were composed of soccer, basketball and baseball players as well as golfers, swimmers and track and field athletes. The two groups (RHI and control) were matched for age, ethnicity and gender. The dataset will be further characterized in the methods section.

At various stages of CTE, clinical symptoms may present similarly to other neurodegenerative disorders such as AD, PD and ALS [86]. However, it is unclear to what degree similarities exist between these disorders along the dimension of speech. Concordantly, we were interested in determining if our model trained on the Verus dataset could be re-purposed to detect AD and PD at or above the current state-of-the-art accuracy on certain AD and PD corpora. Successful transfer learning from the Verus dataset to the aforementioned neurodegenerative disorders would suggest that our model was identifying vocal features in the Verus dataset that are related to brain damage. This would in turn support the utility of speech as a biomarker of RHI-BC.

## 2. Proposed Approach

While deep learning approaches can produce state-of-the-art results in speaker recognition and related tasks, they may require significantly more data to be trained on compared to non-deep learning algorithms. Unfortunately, in the field of medicine, domain-specific data is often difficult to obtain, and algorithms must be trained on datasets of sub-optimal size. Therefore, standard practice is to pretrain a deep learning model on a domain non-specific task before fine-tuning on the task of interest [47, 70]. This allows for the learning of lower-level features which may be domain-unspecific in nature [8]. Building on this standard, we adopted a multi-step transfer learning approach, where we first pretrained our ECAPA-TDNN model on a large, domain non-specific dataset and then subsequently fine-tuned on smaller, domain-specific datasets. This approach outlined below can also be visualized in Figure 1.

**Figure 1:**
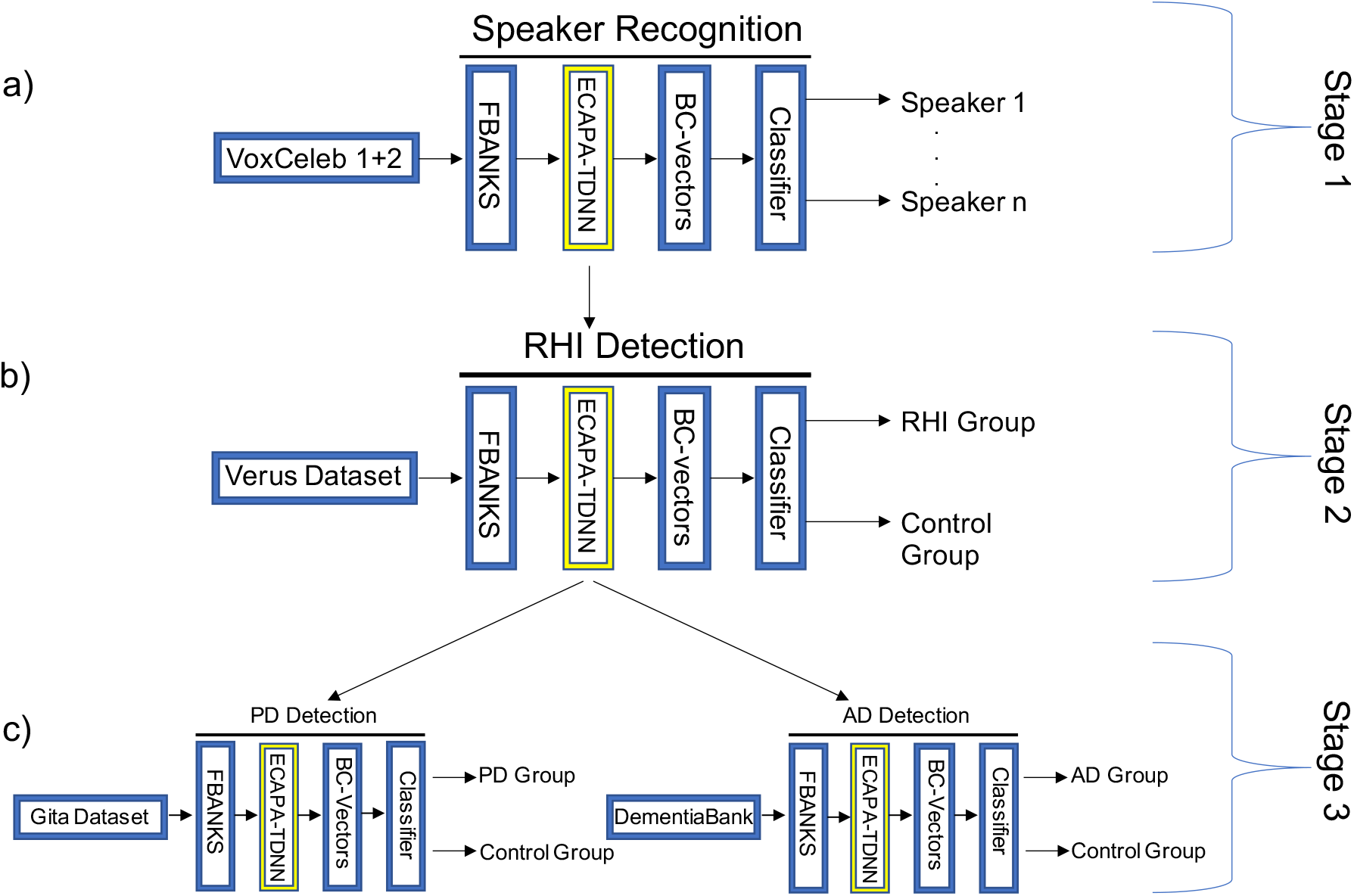
Depiction of the training process used in this study. a) An ECAPA-TDNN model is trained to classify speaker identities with the Voxceleb1+2 dataset. b) The pretrained ECAPA-TDNN model from “a)” is fine-tuned on the Verus dataset to differentiate the RHI from control group. c) Finally, the pre-trained ECAPA-TDNN model from “b)” is fine-tuned on either the DementiaBank or Gita dataset in order to detect AD or PD, respectively.

Stage 1 of our approach entailed training a speaker recognition model. In Stage 2, we used the ECAPA-TDNN model that was pretrained for speaker recognition and fine-tuned it to classify individuals with a history of RHI from those without. In Stage 3, we utilized the ECAPA-TDNN model from Stage 2 and fine-tuned it to perform either AD or PD detection. Using this multi-step transfer learning approach, we start with the broad task of speaker recognition and then successively transfer to the more specific tasks of RHI detection followed by AD and PD detection. This order of operations utilized the larger datasets first, with the intention that the smaller datasets will be used primarily to learn domain-specific features. As some symptoms described in people with a history of RHI present similarly to both PD and AD, it is possible that some of the RHI-specific features learned in Stage 2 will be preserved in Stage 3 [8]. This proposed progression of feature learning is shown in Figure 2.

**Figure 2:**
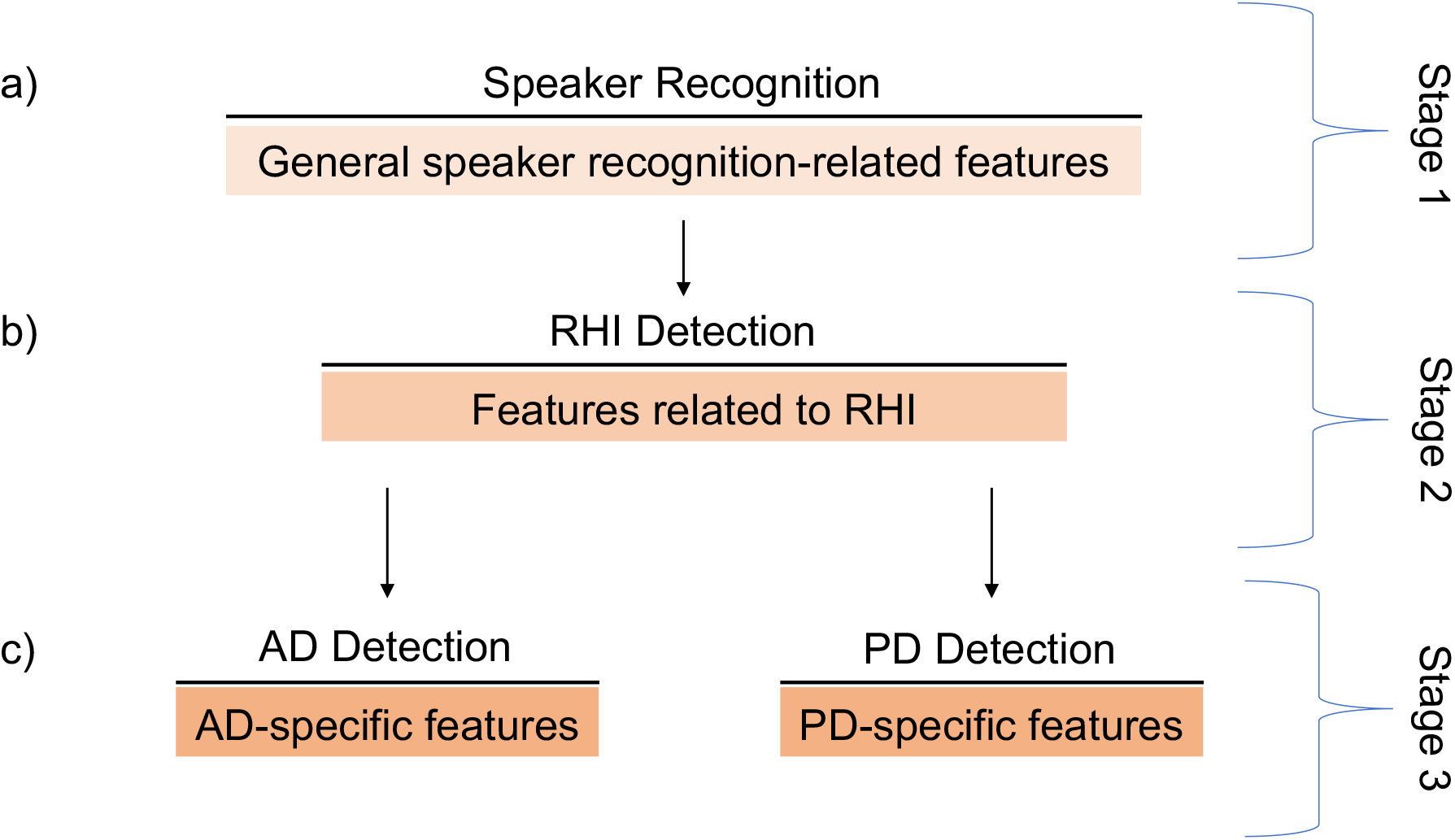
Depiction of the proposed feature learning resulting from the multi-step transfer learning approach. a) Stage 1 results in the learning of low-level features pertinent to speaker recognition. b) Stage 2 involves the transfer of the low-level features from Stage 1 and the learning of RHI-related features. c) Stage 3 involves the transfer of features from Stage 1 and 2 as well as the learning of AD or PD-specific features.

The feature extraction, training and evaluation of the model were mediated through a modified SpeechBrain recipe. Speech-Brain is an opensource toolkit that provides end-to-end methods for various audio processing tasks [67]. The recipe adapted for our project converts raw audio sampled at 16 kHz into short-time Fourier transformed (STFT) filter banks (FBANKS) and augments the data with noise using the Room Impulse Response and Noise (RIRS) Database, while also randomly dropping a specified number of frequencies [45]. FBANKS are widely used features in speech and speaker recognition, especially when using deep learning approaches [24]. The SpeechBrain recipe employs the ECAPA-TDNN model as the embedding extractor. Fundamentally, the ECAPA-TDNN converts variable length utterances into fixed-length embeddings (here termed BC-vectors) which can then be used to characterize speech, as shown in Figure 3. The comparison of BC-vectors determines which speaker (or group) an utterance belongs to. The ECAPA-TDNN model differs from previous TDNN models in the inclusion of ResNet-like layers, termed SE-Res2Blocks, that are interjected with skip connections within the TDNN layers. This allows for greater “… emphasis on channel attention, propagation and aggregation”, which has led to state-of-the-art accuracy in speaker recognition tasks [27, 25]. Once the BC-vectors are extracted, they are output to a classifier which computes the cosine similarity with additive angular margin (AAM) loss [26].

**Figure 3:**
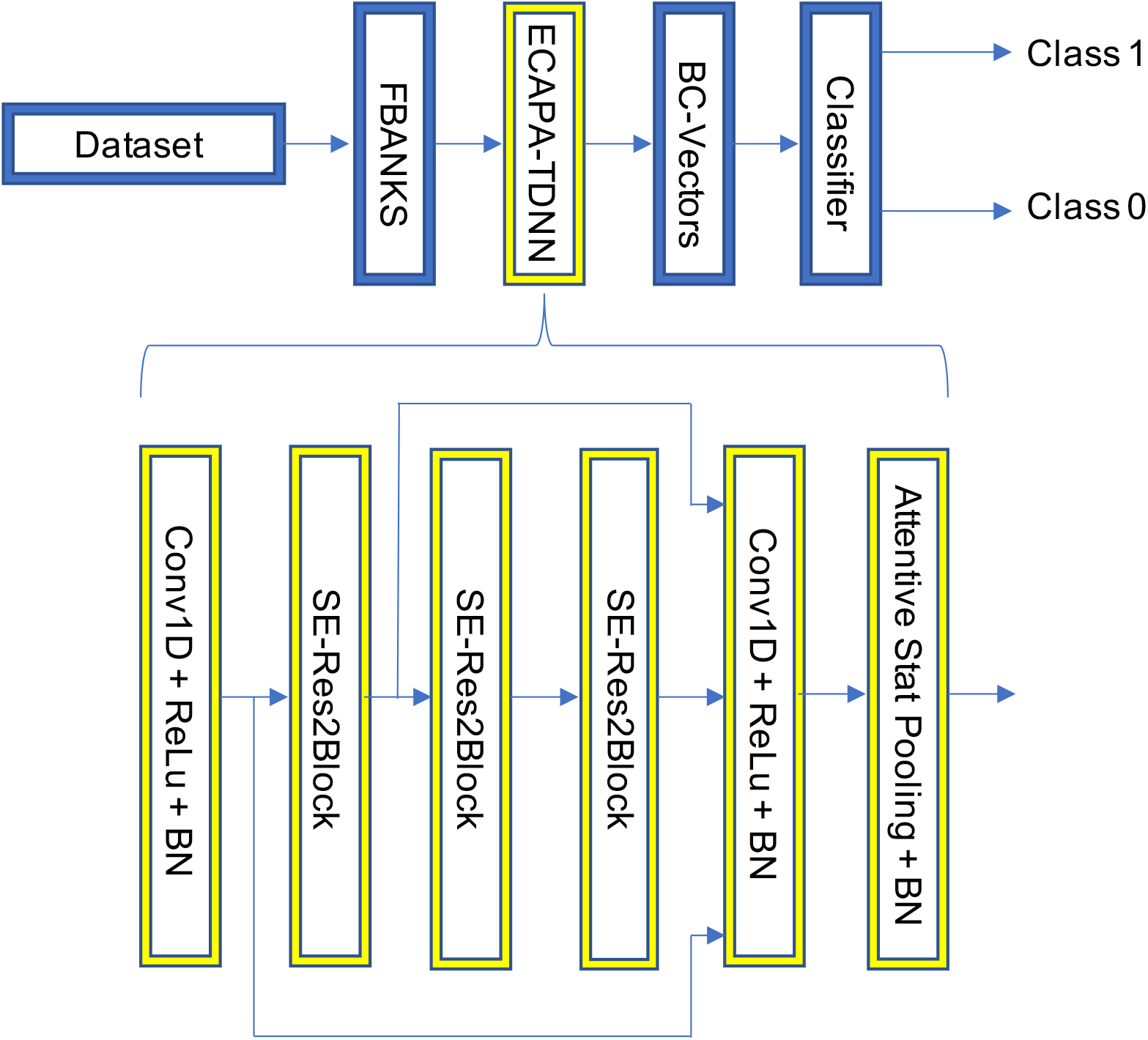
Simplified depiction of the ECAPA-TDNN model used in this study.

## 3. Materials

### 3.1. VoxCeleb Datasets

The VoxCeleb datasets, termed VoxCeleb 1 and VoxCeleb 2 are speech corpora composed of over 2000 hours of speech from over 7000 speakers, downloaded from publicly available videos on YouTube [60]. These datasets are commonly used to benchmark state-of-the-art performance in speaker recognition tasks [19].

### 3.2. Verus Dataset

The Verus dataset is composed of 1210, 13-second recordings, each corresponding to a distinct individual. The recordings were downloaded from publicly available interviews on YouTube and edited using the Audacity application [5]. Each recording is composed of 13 seconds of uninterrupted spontaneous speech. The corpus can be further divided into the RHI and control group, each having 605 recordings. The RHI group is composed of active or retired professional boxers, MMA, Kickboxers and Muay Thai fighters. To be considered a professional, they must have currently or previously competed in a professionally sanctioned league, such as the UFC, Glory, One Championship, Bellator, or other similar leagues. Their affiliation to these leagues was verified by consulting boxrec.com for boxers, glorykickboxing.com for kickboxers, sherdog.com for MMA, and onefc.com for Muay Thai fighters. Professional fighters were selected for the RHI group as data indicate they suffer significantly more RHI and concussions than players of contact sports like football or hockey [84, 65, 17]. The control group is composed of 605 recordings of active or retired professional athletes from non-contact sports such as basketball, baseball, soccer, golf, tennis, cricket, track and field and rowing. Professional athletes were chosen as the control group as a means to control for differences that might exist between professional fighters and non-athletes, such as differences in education, socioeconomic status or hormone levels [2, 33]. The RHI and control groups were controlled for age, gender and race/ethnicity. We also calculated the mean fundamental frequency (F0) and mean energy of all the recordings of fighters and athletes, as displayed in Table 3. The F0 is commonly used as a proxy of the average pitch of a recording [37]. We computed the F0 through Librosa’s use of the YIN algorithm [53]. The mean energy is used to indicate the average intensity or loudness of a recording [44]. The mean energy was also computed using Librosa. These metrics were calculated to ensure that simple, uninformative differences such as pitch or loudness did not exist between the two groups. Tables 1 and 2 provide a more thorough description of the demographic data pertaining to the Verus dataset while Table 3 contains the F0 and mean energy scores. This dataset was used to train the RHI-detection model in Stage 2. Before training, the Verus dataset was split into training and validation sets using stratified k-10 cross validation (CV). The final accuracy will be the average accuracy across the 10 validation folds, with each fold composed of a distinct subset of the data [78]. This CV approach was also applied to the DementiaBank and Gita datasets described below.

**Table 1.** Demographic data of the Verus dataset. * Denotes overlap between “Oceania” and “White”. STD = standard deviation, N = number of subjects.

|  | Control | RHI |
| --- | --- | --- |
| N Total | 605 | 605 |
| N Male | 555 (92%) | 555 (92%) |
| N Female | 50 (8%) | 50 (8%) |
| N Black | 209 (34%) | 208 (34%) |
| N White | 228 (38%) | 226 (37%) |
| N Latin | 72 (12%) | 70 (12%) |
| N Oceania | 60* (10%) | 56* (9%) |
| Mean Age | 35.6 | 36.34 |
| Age STD | 14.21 | 10.22 |

**Table 2.** RHI group of the RHI Control dataset and their corresponding RHI-related history.

| RHI Group | Profession | RHI-Related History |
| --- | --- | --- |
| Terry Norris | Boxer | Boxing license not renewed in year 2000 due to changes in speech and suspicion of suffering from "dementia pugilistica" [64]. |
| James Te Huna | MMA | Retired from the UFC due to brain imaging showing multiple "lesions" and having memory problems [75]. |
| Stephan Bonnar | MMA | Retired from MMA after an MRI showed a cavum septum pellucidum [80]. |
| Chris Benoit | Professional Wrestler | Diagnosed with CTE [38]. |
| Micky Ward | Boxer | Complains of chronic headaches and nausea [20]. |

**Table 3.** Demographic data within the control and RHI groups of the Verus dataset. * Denotes overlap between “Oceania” and “White”. STD = standard deviation, N = number of subjects.

|  | Control |  |  |  | RHI |  |
| --- | --- | --- | --- | --- | --- | --- |
| Sport | Baseball | Basketball | Golf | Soccer | Boxing | MMA |
| N Total | 187 | 186 | 46 | 104 | 283 | 307 |
| N Black | 28 (15%) | 147 (80%) | 1 (3%) | 24 (23%) | 126 (48%) | 72 (25%) |
| N White | 114 (61%) | 22 (12%) | 38 (83%) | 56* (54%) | 80* (31%) | 151* (53%) |
| N Latin | 40 (21%) | 10 (5%) | 3 (7%) | 12 (12%) | 27 (10%) | 36 (13%) |
| N OCE (Oceania) | 5 (2.6%) | 1 (0.5%) | 3* (7%) | 33* (32%) | 51* (20%) | 5* (2%) |

### 3.3. RHI Control Dataset

The RHI control dataset consists of 10, 13 second voice recordings. The recordings were downloaded from publicly available interviews on YouTube and edited using the Audacity application [5]. The dataset is divided into a “RHI” and “control” group. The RHI group consists of 5 individuals who have a history of either CTE, probable CTE, or RHI-BC. The control group consists of three chess players and two golfers. The relevant medical history of the RHI group is shown in Table 2.

### 3.4. DementiaBank Dataset

The DementiaBank dataset was created by [7] as a means to better characterize the behavioural neurological factors that can aide in diagnosing AD. The section of this dataset used in our study was the “Cookie Theft” description task, where a subject was asked to describe a scene shown as a picture. We included recordings of participants who were listed as either “probable AD” or “possible AD”, resulting in 259 recordings from 167 speakers. We also included 236 recordings from 97 healthy controls. Each recording was edited to include 13 seconds of uninterrupted spontaneous speech, using Audacity [5]. In total, 60 recordings did not contain 13 seconds of uninterrupted speech and were therefore edited to include the maximum amount of uninterrupted speech under 13 seconds.

### 3.5. Gita Dataset

The Gita dataset is a speech corpus consisting of 50 subjects with PD and 50 healthy controls matched for age and gender [62]. The corpus included in this study is that of a monologue, where participants were asked to describe their daily routines. We included 100, 13 second recordings of uninterrupted spontaneous speech of each individual listed in the study. This resulted in 50 recordings of subjects with PD and 50 recordings of healthy controls. Each recording was edited using Audacity [5].

## 4. Results and Discussion

### 4.1. RHI Detection

The average accuracy over 10 folds of the Verus dataset was 85%. The accuracy of each fold, as well as the average accuracy and standard deviation are shown in Table 4. The youngest age of individuals correctly classified to the RHI group was 21, all four of which were boxers. This is the first time we are aware of that speech has been used to accurately differentiate individuals with a history of significant RHI from those without. Speech problems have long been associated with RHI, yet how they relate to RHI-BC or CTE severity has not been sufficiently explored. Literature pertaining to boxers indicates that motor speech disorders occur relatively frequently [51, 23, 46]. However, the prevalence of motor speech disorders in the clinical description of CTE as depicted by [55] is less clear. Dysarthria is sometimes listed as a symptom of CTE, while language impairments are commonly listed as symptoms of advanced stages of CTE [58, 4]. Motor speech disorders differ from language impairments in both their symptoms and underlying pathophysiology. Motor speech disorders (such as dysarthria) are characterized by difficulties in producing or pronouncing words or sounds [28]. Language impairments refer to difficulty in understanding or creating language, which may occur without any difficulties in the mechanical production of sounds [12]. Motor speech disorders are generally caused by damage to regions in the brain responsible for the motor control and coordination of the muscles used to create speech. Regarding professional fighters, brain damage has been found in the form of cerebellar scarring, depigmentation of the substantia nigra, pyramidal and extrapyramidal dysfunction and cerebral atrophy [17, 21, 58]. Cerebellar scarring can lead to ataxic dysarthria, a condition which presents with irregular rate of speech and stress patterns, scanning speech and improper articulation [74]. Extrapyramidal dysfunction and depigmentation of the substantia nigra can result in parkinsonism, a condition where speech may be affected by changes in speed, decreased volume and pitch variability, pronunciation abnormalities and decreased intelligibility [88, 35]. Indeed, parkinsonism, ataxic dysarthria and other forms of dysarthria have all been documented in professional fighters, while parkinsonism is also listed as a symptom of advanced CTE [55, 17, 21, 58]. However, language impairments can be caused by damage to cortical regions of the brain [12]. Their occurrence in the later stages of CTE is likely due to the pathological progression of CTE, where cortical regions tend to be affected later in the disease [1]. Further, [9] found that although significant differences in regional brain volumes existed between professional fighters and controls, cognitive tests were largely indistinguishable. These data seem to indicate that higher order processes like language and cognition are not obviously affected in the early stages of CTE or RHI-BC. This has been indicated in other neurodegenerative disorders [36, 41, 83]. However, motor speech disorders, which are caused by damage to subcortical or hindbrain regions, might manifest significantly earlier. This would be consistent with our finding that four boxers at just 21 years of age were correctly classified to the RHI group.

**Table 4.**
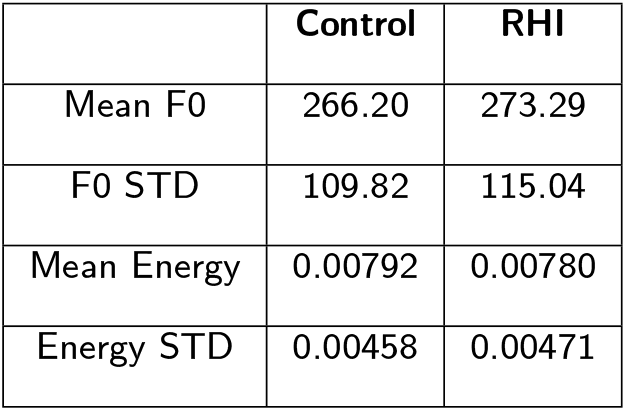
Average F0 and mean energy scores calculated using the Verus dataset. STD = standard deviation.

|  | Control | RHI |
| --- | --- | --- |
| Mean F0 | 266.20 | 273.29 |
| F0 STD | 109.82 | 115.04 |
| Mean Energy | 0.00792 | 0.00780 |
| Energy STD | 0.00458 | 0.00471 |

### 4.2. RHI Control

A limitation of the relatively large Verus dataset is that we could not gather relevant medical history of every individual. As not every professional fighter will develop a neurodegenerative disorder, and some athletes in non-contact sports can indeed develop neurodegenerative disorders [48], we reasoned that the accuracy of our model on the Verus dataset might be lower than in a clinically controlled setting. Thus, we created a small dataset in which the RHI group was composed of individuals who were either diagnosed with CTE, suspected of having CTE, or had received brain imaging results suggestive of trauma-induced damage. Further, the control group was composed of three chess players (i.e. non-athletes) and 2 golfers. The relevant information of these individuals is shown in Table 5. Our model trained on the Verus dataset achieved 100% accuracy in predicting whether an individual belonged to the RHI or control group. While this is a small sample size, there are some points that merit consideration. Our model accurately classified an individual who was diagnosed post-mortem as having CTE, but was a professional wrestler, not a fighter. Further, our model also accurately classified the three chess players as belonging to the control group. This suggests the model can generalize to individuals who were not either professional fighters nor athletes. These findings also suggest that our model may perform at over 85% accuracy when the medical history of the subjects is already known.

**Table 5.**
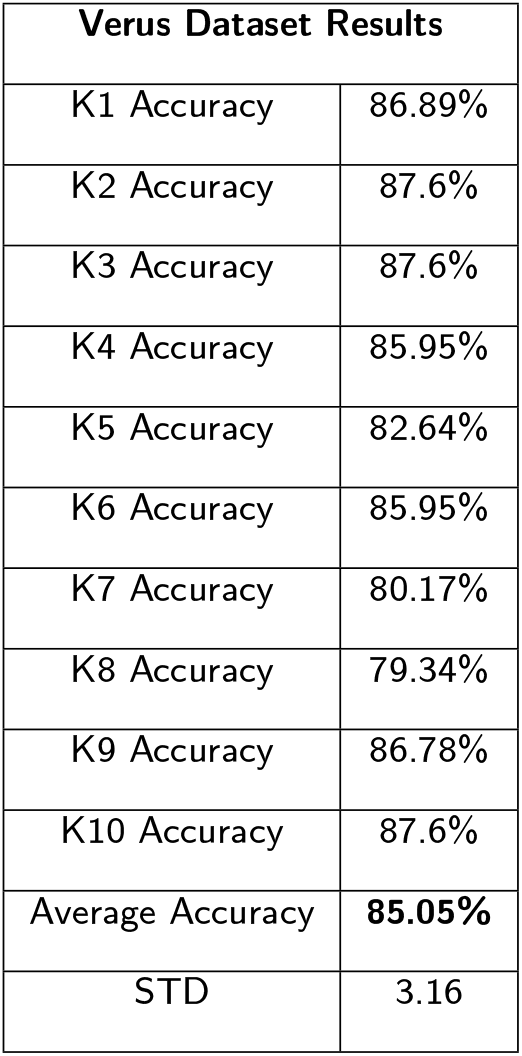
K-Fold Cross Validation Results of the Verus Dataset. STD = standard deviation

### 4.3. Alzheimer’s Disease Detection

The average accuracy over 10 folds of the DementiaBank dataset was 84.99%, as shown in Table 6. This is the highest accuracy we are aware of achieved on this dataset. [29] achieved 81.9% accuracy using logistic regression and what was described as a mixture of 4 latent variables, generally corresponding to syntax/fluency, semantics, acoustics, and other factors. In a more recent publication, [90] achieved F1-micro and F1-macro scores of 0.799. They again used a combination of acoustic and linguistic features, with “Consensus Networks” (CN), deep neural networks that contain a “discriminator” which functions similarly to a generative adversarial network (GAN). Further, [89] achieved 88% accuracy on a subset of the DementiaBank dataset, while [40] achieved 91% accuracy. However, ([89] allowed for “spillover”, meaning data from the same patients were used in both the training and validation set. Further, [40] included transcripts from at least the “Cookie Theft” and “Recall” tasks and did not indicate if they also included transcripts from the “Sentence” and “Fluency” tasks. They also did not indicate whether they included patients diagnosed with a disorder other than AD in the AD group. Nonetheless, the literature indicates that attempts at using machine learning to detect AD rely more on linguistic than acoustic features [49]. This trend might exist due to theoretical knowledge pertaining to AD. Specifically, the symptoms of AD are often attributed to the accumulation of misfolded proteins in the cerebral cortex. It is thought that memory and cognition problems are amongst the first symptoms in AD, with motor problems not occurring until later stages when the misfolded proteins have significantly accumulated in subcortical structures [87]. However, recent findings have shown that motor problems are prevalent in AD and might present preclinically [15, 79, 14]. This aligns with our findings, as we achieved state-of-the-art performance on the DementiaBank dataset using a model which is text independent. To clarify, this means our model was not able to utilize or learn any linguistic features, but instead was able to differentiate subjects based on speech characteristics like syllable rate, pitch and others. Our findings provide motivation to further investigate the motor changes evident in speech which could serve as an early biomarker of AD.

**Table 6.** K-Fold Cross Validation Results of the DementiaBank dataset. STD = standard deviation

| DementiaBank Results |  |
| --- | --- |
| K1 Accuracy | 91.67% |
| K2 Accuracy | 91.67% |
| K3 Accuracy | 89.6% |
| K4 Accuracy | 77.1% |
| K5 Accuracy | 81.2% |
| K6 Accuracy | 87.5% |
| K7 Accuracy | 87.5% |
| K8 Accuracy | 81.2% |
| K9 Accuracy | 77.1% |
| K10 Accuracy | 85.4% |
| Average Accuracy | **84.99%** |
| STD | 5.54 |

### 4.4. Parkinson’s Disease Detection

The average accuracy over 10 folds of the Gita dataset was 89% with a standard deviation of 7.3, as shown in Table 7. The accuracy reported in [62] on this section of the dataset was 81% with a standard deviation of 7. To determine if the difference between these two scores was statistically significant, we performed a two-tailed t-test assuming unequal variance and an alpha value of 0.05. As the t-test indicated a p-value of 0.022, we can assume our higher accuracy is statistically significant from that achieved by [62].

**Table 7.** K-Fold Cross Validation Results of the Gita Dataset. STD = standard deviation

| Gita Dataset Results |  |
| --- | --- |
| K1 Accuracy | 90% |
| K2 Accuracy | 90% |
| K3 Accuracy | 100% |
| K4 Accuracy | 90% |
| K5 Accuracy | 80% |
| K6 Accuracy | 80% |
| K7 Accuracy | 80% |
| K8 Accuracy | 100% |
| K9 Accuracy | 90% |
| K10 Accuracy | 90% |
| Average Accuracy | 89% |
| STD | 7.37 |

## 5. Limitations

As previously stated, the current trend in speaker recognition and related tasks is to use deep learning approaches which learn features, as opposed to the previously standard approach of engineering features to be fed to algorithms which do not use deep neural networks [39]. This has lead to a limitation in our study in the form of a trade-off. Namely, our deep learning approach results in higher accuracy at the expense of interpretability. However, as the relationship between speech and RHI-BC has not been sufficiently characterized, we reasoned feature engineering would be inferior to our deep learning approach as it is not known which features are relevant to the task. Another limitation to our study is that the medical history of the subjects used was not known. We have attempted to address this limitation, yet further studies in a clinically controlled setting are needed. Another limitation was that we did not control for bilingualism. Recent data indicate that bilingualism may act as a form of “cognitive reserve”, thus delaying onset of symptoms in neurodegenerative disorders like AD [56, 22]. It should be noted that bilingualism appears to delay deficits in executive functioning and has not been linked to motor-speech disorders, yet it is still possible that our classification accuracies would be higher if we excluded individuals who were bilingual.

## 6. Conclusion

The long-term consequences of RHI are still being investigated, with indications that a history of RHI can increase the risk of developing CTE, AD, ALS, PD and other disorders [13, 34, 43, 42, 76]. Unfortunately, there are no accepted biomarkers of RHI-BC or CTE, thus tracking their progression and mapping the level of RHI exposure to disease severity has proven difficult [9]. In this study, we found that a 13 second voice recording can be used to differentiate those with a history of RHI from this without at 85% accuracy. We then used the ECAPA-TDNN model trained on RHI detection to achieve state-of-the-art results in detecting AD and PD on the DementiaBank and Gita datasets, respectively. Machine learning techniques have often focused heavily on language impairments when detecting AD [29, 40, 49, 63]. Our study is the first we are aware of to focus exclusively on acoustic, text-independent features of speech. This might indicate that motor speech problems can serve as a biomarker of AD. Further, our success in transfer learning is evidence that the features learned by the ECAPA-TDNN model trained on the Verus dataset were related to speech changes associated with brain damage. Finding a biomarker of RHI-BC or CTE that presents early in disease progression could help with risk management and improve patient outcome. Indeed, our model accurately classified 4 boxers of just 21 years of age as belonging to the RHI group, suggesting speech might serve as a such a biomarker. Future research should aim to characterize the relationship between speech disorders and progression of RHI-related neurodegenerative disorders.

## Data Availability

All code is available upon request. Externally owned data may be available through the provided URLs. The Verus dataset will be available upon request and following copyright registration.

https://dementia.talkbank.org/

https://www.robots.ox.ac.uk/~vgg/data/voxceleb/

## 7. Acknowledgements

The authors would like to thank Dr. Mariano Avino for his valuable support and assistance.

